# COVID-19 in children in NSW, Australia, during the 2021 Delta outbreak: Severity and Disease spectrum

**DOI:** 10.1101/2021.12.27.21268348

**Authors:** Phoebe Williams, Archana Koirala, Gemma Saravanos, Laura Lopez, Catherine Glover, Ketaki Sharma, Tracey Williams, Emma Carey, Nadine Shaw, Emma Dickins, Neela Sitaram, Joanne Ging, Paula Bray, Nigel Crawford, Brendan McMullan, Kristine Macartney, Nicholas Wood, Beth Fulton, Christine Lau, Philip N Britton

## Abstract

**Objective(s):** To describe the severity and clinical spectrum of SARS-CoV-2 infection in Australian children during the 2021 Delta outbreak.

**Design, Setting & Participants:** A prospective cohort study of children <16 years with a positive SARS-CoV-2 nucleic acid test cared for by the Sydney Children’s Hospital Network (SCHN) virtual and inpatient medical teams between 1 June – 31 October 2021.

**Main outcome measures:** Demographic and clinical data from all admitted patients and a random sample of outpatients managed under the SCHN virtual care team were analysed to identify risk factors for admission to hospital.

**Results:** There were 17,474 SARS-CoV-2 infections in children <16 years in NSW during the study period, of whom 11,985 (68.6%) received care coordinated by SCHN. Twenty one percent of children infected with SARS-CoV-2 were asymptomatic. For every 100 SARS-CoV-2 infections in children <16 years, 1.26 (95% CI 1.06 to 1.46) required hospital admission for medical care; while 2.46 (95% CI 2.18 to 2.73) required admission for social reasons only. Risk factors for hospitalisation for medical care included age <6 months, a history of prematurity, age 12 to <16 years, and a history of medical comorbidities (aOR 7.23 [95% CI 2.92 to 19.4]). Of 17,474 infections, 15 children (median age 12.8years) required ICU admission; and 294 children required hospital admission due to social or welfare reasons.

**Conclusion:** The majority of children with SARS-CoV-2 infection (Delta variant) had asymptomatic or mild disease. Hospitalisation was uncommon and occurred most frequently in young infants and adolescents with comorbidities. More children were hospitalised for social reasons than for medical care.

## BACKGROUND

Since the ancestral SARS-CoV-2 virus emerged in Wuhan in December 2019, Coronavirus Diseases (COVID-19) has caused half a billion deaths worldwide; yet only a very small minority of these have occurred in children.^1,2^ Whilst limited paediatric specific seroprevalence data limit precise estimation, it is clear that mortality due to SARS-CoV-2 in children is extremely rare, with an estimated infection fatality rate (IFR) of 5 per 100,000 cases in people under 18 years.^3^ Similarly, hospitalisation in children is also uncommon,^4–7^ and SARS-CoV-2 infection has been consistently described to be asymptomatic or cause mild disease in most children.^5,6,8–16^ However, there are limited data tracking the true severity and clinical spectrum of COVID-19 in children, particularly with regard to the variant of concern (VOC) B1.617.2, also known as the ‘Delta’ VOC.^17^

Most published reports on COVID-19 symptomatology in children describe hospitalised cohorts infected with the ancestral strain or earlier VOCs, and there are few data describing children with SARS-CoV-2 infection who are well enough to be managed in the community.^9,17,18^ Moreover, hospitalisation data regarding children with SARS-CoV-2 infection frequently fails to delineate the causes of hospitalisation, which may provide important detail as to the true spectrum of illness in this population. Two single centre studies from the USA revealed that almost half of all paediatric admissions with SARS-CoV-2 were incidental infections in children admitted due to other diagnoses;^19,20^ while 21% of paediatric cases in a large UK multicentre study were similarly found to be admitted to hospital with, rather than due to, SARS-CoV-2 infection.^5^

Australia curtailed the spread of COVID-19 in 2020 through aggressive public health suppression strategies utilising non-pharmaceutical interventions and travel restrictions.^21–23^ Seroprevalence in the paediatric population to March 2021 was subsequently very low, estimated at 0.23% (95% CI 0.06 to 0.57%), suggesting that a majority of the paediatric population had not been infected with SARS-CoV-2 (Koirala *et al*., Seroprevalence of SARS-CoV-2 specific antibodies in Australian children between Nov 2020 and Mar 2021; MJA, unpublished).

In June 2021, an outbreak of the SARS-CoV-2 Delta VOC arose from a single case in New South Wales (NSW), the most populous state in Australia (population 8.1 million).^24^ Public health measures such as school closures were promptly enforced to reduce community transmission and shield children (who were largely unvaccinated), at considerable cost to their education, wellbeing and development.^25,26^

SARS-CoV-2 infection may result in asymptomatic infection or a wide spectrum of clinical lower respiratory tract disease (COVID-19) in adults, and this has been well characterised. By contrast, clinical features and risk factors for severe disease with SARS-CoV-2 infection in children are less well described.^6^ We aimed to describe the severity and clinical spectrum of SARS-CoV-2 infection in a prospective cohort of children managed through community-based virtual care and hospital admission during the 2021 Delta outbreak in NSW, Australia, to determine hospitalisation rates for both medical and social indications in children infected with SARS-CoV-2 and identify risk factors for severe disease.

## METHODS

### Clinical Setting & Data Sources

The Sydney Children’s Hospital Network (SCHN) virtualKIDS COVID-19 positive outpatient response team (virtualKIDS-CORT, hereafter referred to as virtual care) service was established in 2020, and provides comprehensive, community-based virtual care for children with SARS-CoV-2 infection in metropolitan Sydney. Based on a Hospital-in-the-Home (HITH) platform, the service provides telehealth and outreach in-home clinical assessments by a medical officer and nursing team. The service cared for 90 children in 2020 and was rapidly scaled-up from June 2021 to support increasing cases of SARS-CoV-2 infection in children across Greater Sydney during the Delta outbreak.

All positive SARS-CoV-2 nucleic acid test (NAT) and serology results in children <16 years in NSW are recorded in the Notifiable Conditions Information Management System (NCIMS). Close contacts of positive cases must isolate and undergo SARS-CoV-2 NAT via real time polymerase chain reaction (RT-PCR) prior to release from quarantine, identifying additional SARS-CoV-2 positive children who may be asymptomatic. Positive results trigger a public health response comprising of a case interview and contact tracing.

SARS-CoV-2 positive children living in three local health districts (all-age population: 3 million) were routinely referred to the virtual care service for symptom and welfare review. Clinical and social data are collected via regular (twice daily to twice weekly, based on the clinical and social triage risk) telehealth reviews; or in-person home medical reviews where necessary.

Children admitted to hospital had further clinical data obtained via the Paediatric Active Enhanced Disease Surveillance (PAEDS, www.paeds.org.au) surveillance teams, a national collaboration across Australia’s tertiary children’s hospitals. Children may be admitted for either medical reasons (that is, requiring medical care due to SARS-CoV-2 infection or due to an alternative diagnosis, with incidental SARS-CoV-2 infection); or as ‘social’ admissions – encompassing children admitted due to parents and carers requiring hospitalisation due to COVID-19, children admitted under public health orders, or children with chronic issues whose usual support networks were unable to care for them in the context of a SARS-CoV-2 positive status (including children in out-of-home-care, children under the statutory child protection care of the Department of Communities and Justice [DCJ], and those requiring mental health support).

PAEDS surveillance nurses actively reviewed clinical and laboratory records of children with SARS-CoV-2 infection or paediatric inflammatory multi-system syndrome temporally associated with SARS-CoV-2 (PIMS-TS, also known as multi-system inflammatory syndrome in children, MIS-C) and liaised with clinical teams to identify and subsequently monitor eligible children presenting to participating hospitals, resulting in comprehensive, individual case data collated within a national electronic database (REDCap, hosted at the University of Sydney) that facilitated analyses and reporting.

### Analysis

We described age-specific childhood frequencies of notified SARS-CoV-2 infections in NSW as recorded in NCIMS, virtual care outpatients and SCHN inpatients in children aged <16 years by epidemiologic week between June 1 and October 31, 2021. We calculated a COVID-19 medical admission rate and COVID-19 social admission rate using SCHN virtual care admissions as a denominator, with 95% confidence intervals; and a COVID-19 ICU admission rate and PIMS-TS admission rate using NSW-wide NCIMS notifications infections as a denominator (Supplementary methods).

We described patient demographics, medical comorbidities, clinical features, diagnosis, treatment, and outcomes (non-intensive care hospital admission, intensive care admission, death) among all children admitted to the two SCHN hospitals for medical and non-medical reasons. Primary clinical manifestations or clinician diagnosis for admitted cases was collected from discharge summaries.

We identified a simple random sample of children cared for in the community by the SCHN virtual care service (stratified by calendar month; sampled to achieve 2:1 ratio of ambulatory to admitted cases). Clinical data from this sample was combined with inpatient data from the two SCHN hospitals, obtained via the PAEDS network. We analysed data for associations between demographics, medical comorbidities and clinical features comparing the admitted cases and the sample of virtual care cases in a univariate and multivariate logistic regression model. Given an absence of measured or self-reported weight amongst virtual care children, we imputed to these cases the highest population prevalence estimate (28.9%) for overweight amongst NSW children.^27^

### Approvals

PAEDS surveillance is approved by the SCHN human research ethics committee (HREC; study number 2019/ETH06144) including a waiver of consent for collection of case data. The aggregate reporting of SCHN virtual care data and NSW wide NCIMS data was approved by the NSW Ministry of Health and SCHN.

## RESULTS

Between June 1 - October 31 2021 there were 17,474 confirmed SARS-CoV-2 infections in children aged 0 to <16years in NSW, of whom 11,985 (68.6%) received virtual care coordinated by SCHN (Figure 1). A total of 459 children were admitted to SCHN hospitals; 165 (35.9%) were admitted due to medical need (Table 1) and 294 (64.1%) were admitted for social reasons (Table 2). The age distribution of NSW SARS-CoV-2 infections and virtual care admissions was similar, but medical admissions were more common in children aged <2 years, and ICU admissions in children aged 12-15 years. 15 children were admitted to ICU, of whom 60% were aged ≥12 years (Table 3).

**Figure 1.**
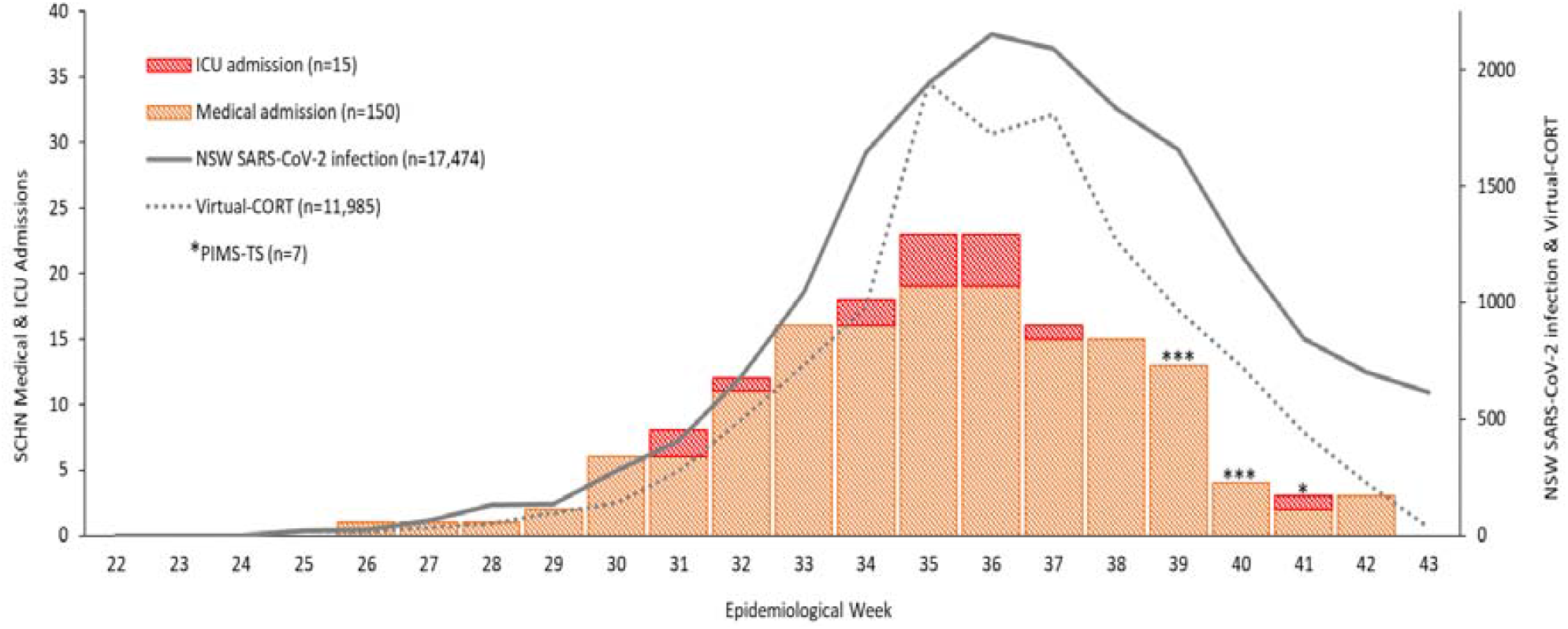
Epidemiologic curve of COVID-19 in children aged 0-15 years during the NSW Delta outbreak, 2021 (June 1 - October 31) Notes: NSW SARS-CoV-2 infections in children aged 0-15 years by earliest confirmed date; children admitted to VirtualKIDS- CORT aged 0 to <16 years by admitted date; children recruited to PAEDS COVID-19 surveillance, admitted to Children’s Hospital at Westmead and Sydney Children’s Hospital, Randwick (Sydney Children’s Hospital Network [SCHN]) for medical reasons and aged 0 to <16 years by laboratory confirmation of SARS-CoV-2, children recruited to PAEDS PIMS-TS surveillance aged 0 to <16 years by date of illness onset.

**Table 1:** Age stratified case frequency of SARS-CoV-2 infections in children aged 0-15 years in NSW and managed through SCHN, June 1 - October 31, 2021.

| Age Group | NSW | % | Virtual KIDS-CORT | % | Medical Admit | % | ICU Admit | % | PIMS-TS | % |
| --- | --- | --- | --- | --- | --- | --- | --- | --- | --- | --- |
| **0-5 months** | 510 | 2.9 | 416 | 3.5 | 49 | 33.1 | 1 | 6.3 | 1 | 14.3 |
| **6-23 months** | 1,612 | 9.2 | 1,154 | 9.6 | 26 | 17.2 | 1 | 6.3 | 1 | 14.3 |
| **2-4 years** | 3,289 | 18.8 | 2,307 | 19.2 | 20 | 13.2 | 0 | 6.3 | 3 | 42.9 |
| **5-11 years** | 7,685 | 44.0 | 5,076 | 42.4 | 22 | 14.6 | 4 | 25.0 | 2 | 28.6 |
| **12-15 years** | 4,378 | 25.1 | 3,032 | 25.3 | 33 | 21.9 | 9 | 56.3 | 0 | 0 |
| **TOTAL** | **17,474** | **100.0** | **11,985** | **100.0** | **150** | **100.0** | **15** | **100.0** | **7** | **100.0** |
| **Median Age (IQR)** | 7.9<br>(4.1-12.0) |  | 7.9<br>(3.9-12.1) |  | 2.0<br>(0.3-11.2) |  | 12.8<br>(10.4-15.5) |  | 3.2<br>(1.8-7.5) |  |
| **Rate per 100 SARS-CoV-2 infections (95% CI if applicable)** |  |  | N/A |  | 1.26*<br>[1.06 - 1.46] |  | 0.09^ |  | 0.04^ |  |

Abbreviations: ICU=intensive care unit; IQR=inter-quartile range; NSW=New South Wales; PIMS-TS=paediatric inflammatory multi-system syndrome temporally associated with SARS-CoV-2 infection.
\*VirtualKIDS-CORT cases used as denominator. ^NSW-wide notifications used as denominator with PIMS-TS = admission rate calculated using data cut of at 30 September, 2021 given lag time for this 'post-infectious' clinical manifestation.

**Table 2:** Demographic data and reason for admission among for children aged 0-18 years admitted to SCHN for social (non-medical) reasons – these are also known as ‘Home in the Hospital’ admissions.

| **Social admissions** | **N** |
| --- | --- |
| Total number of admitted CYP | 294 |
| Admission rate per 100 SARS-CoV-2 infections (95% CI)* | 2.45 [2.18 - 2.73] |
| Total number of family units | 162 |
| Median age (yrs; IQR) | 8.00 (4.00; 12.75) |
| Median length of stay (days; IQR) | 5 (3; 10) |
| **Admission by age groups (yrs)** | **n (%)** |
| <2 | 41 (14) |
| 2-4 | 45 (15) |
| 5-11 | 117 (40) |
| 12-15 | 84 (29) |
| 16-18 | 7 (2) |
| **Initial reason for admission** | **n (%)** |
| Parent/carer impacted by COVID-19** | 202 (69) |
| CYP admitted under Public Health order | 5 (2) |
| COVID-19 diagnosis impacted CYP's care and/or safety |  |
| DCJ/OOHC supports impacted | 30 (10) |
| Disability supports impacted | 7 (2) |
| Mental health / behaviour impacted | 6 (2) |
| Other |  |
| Unaccompanied minor for medical admission | 34 (12) |
| Boarder sibling | 10 (3) |

Abbreviations: CYP=child and young person; DCJ=department of communities and justice, NSW; IQR=inter-quartile range; OOHC=out of home care; yrs=years
\*VirtualKIDS-CORT cases used as denominator.
\*\*Includes families in which both parents were admitted to hospital due to acute COVID-19, and families in which one parent was admitted with COVID-19 while the remaining carer was unable to support / care for all other children within the household alone (potentially also unwell with mild COVID-19).

**Table 3.** Demographics and comorbid conditions of patients with SARS CoV-2 admitted to ambulatory care, medical (non-ICU) care and ICU at SCHN hospitals June - October 2021.

| Characteristic | VirtualKIDS-CORT<br>sample,<br>N = 344<sup>1</sup> | Medical (non-ICU)<br>admission,<br>N = 150<sup>1</sup> | ICU admission,<br>N = 15<sup>1</sup> | P value<sup>2</sup> |
| --- | --- | --- | --- | --- |
| **Median age<sup>1,3</sup>** | 7.8 (3.8, 11.4) | 2.0 (0.3, 11.2) | 12.8 (10.4, 15.5) | **<0.001<sup>11</sup>** |
| **Age group<sup>3</sup>** |  |  |  |  |
| <6 months | 13 / 344 (3.8%) | 49 / 150 (33%) | 1 / 15 (6.7%) | **<0.001<sup>11</sup>** |
| 6 months to <2 years | 34 / 344 (9.9%) | 26 / 150 (17%) | 1 / 15 (6.7%) | 0.060 |
| 2 to <5 years | 67 / 344 (19%) | 20 / 150 (13%) | 0 / 15 (0%) | **0.048** |
| 5 to <12 years | 157 / 344 (46%) | 22 / 150 (15%) | 4 / 15 (27%) | **<0.001<sup>11</sup>** |
| 12 to <16 years | 73 / 344 (21%) | 33 / 150 (22%) | 9 / 15 (60%) | **0.005<sup>11</sup>** |
| **Sex (M)** | 189 / 344 (55%) | 78 / 150 (52%) | 9 / 15 (60%) | 0.8 |
| **Indigenous<sup>4</sup>** | 13 / 334 (3.9%) | 8 / 147 (5.4%) | 1 / 15 (6.7%) | 0.4 |
| **Vaccination status<sup>5</sup>** |  |  |  | 0.4 |
| Unvaccinated | 313 / 321 (98%) | 135 / 137 (99%) | 12 / 13 (92%) |  |
| 1 Dose | 5 / 321 (1.6%) | 2 / 137 (1.5%) | 1 / 13 (7.7%) |  |
| 2 Doses | 3 / 321 (0.9%) | 0 / 137 (0%) | 0 / 13 (0%) |  |
| **Household contact<sup>6</sup>** | 285 / 306 (93%) | 128 / 138 (93%) | 12 / 14 (86%) | 0.4 |
| **Prematurity (≤ 36 weeks gestation)** | 13 / 209 (6.2%) | 17 / 123 (14%) | 2 / 13 (15%) | **0.037** |
| **Weight > 95th percentile<sup>7</sup>** | 99 / 344 (29%) | 27 / 139 (19%) | 8 / 14 (57%) | **0.004<sup>11</sup>** |
| **Comorbid condition<sup>8</sup>** | 62 / 337 (18%) | 52 / 149 (35%) | 9 / 15 (60%) | **<0.001<sup>11</sup>** |
| Asthma<sup>9</sup> | 35 / 62 (56%) | 13 / 52 (25%) | 3 / 9 (33%) | **0.002<sup>11</sup>** |
| VIW/recurrent wheeze<sup>9</sup> | 0 / 62 (0%) | 6 / 52 (12%) | 0 / 9 (0%) | **0.017** |
| Other respiratory disease<sup>9</sup> | 1 / 62 (1.6%) | 7 / 52 (13%) | 1 / 9 (11%) | **0.038** |
| Cardiac disease | 4 / 62 (6.5%) | 6 / 52 (12%) | 2 / 9 (22%) | 0.2 |
| Neurological disease | 3 / 61 (4.9%) | 5 / 52 (9.6%) | 1 / 9 (11%) | 0.5 |
| Immunodeficiency | 1 / 62 (1.6%) | 3 / 52 (5.8%) | 0 / 9 (0%) | 0.5 |
| Renal disease | 0 / 62 (0%) | 1 / 51 (2.0%) | 1 / 9 (11%) | 0.067 |
| Liver disease | 0 / 62 (0%) | 2 / 52 (3.8%) | 0 / 9 (0%) | 0.3 |
| Diabetes | 0 / 62 (0%) | 1 / 52 (1.9%) | 0 / 8 (0%) | 0.5 |
| Other<sup>10</sup> | 24 / 62 (39%) | 32 / 52 (62%) | 7 / 9 (78%) | **0.013<sup>11</sup>** |

Boldfaced P values are significant at an alpha level of 0.05. Abbreviations: ICU, intensive care unit; VIW, viral induced wheeze. Numerator and denominator data source is PAEDS. Denominators less than the sample size (N) are due to missing data.
<sup>1</sup>Median (IQR); n / N (%); <sup>2</sup>Kruskal-Wallis rank sum test; Fisher's exact test; Pearson's Chi-squared test; <sup>3</sup>Age at confirmation of positive PCR test; <sup>4</sup>Aboriginal and Torres Strait Islander child; <sup>5</sup>Dose 1 dates for 2 ambulatory care cases unconfirmed, case excluded if dose 1 date after positive covid PCR test confirmation; <sup>6</sup>Defined as a confirmed case that was a household contact; <sup>7</sup>Presumed correlate of obesity obtained for Ambulatory care patients based on 28.9% population prevalence (NSW Ministry of Health, 2020); <sup>8</sup>Medical history was reviewed and records consistent with a medical comorbidity were reported; <sup>9</sup>Total number of respiratory diseases: asthma (n = 51), VIW (n = 5), recurrent wheeze (n = 1); <sup>10</sup>Other respiratory diseases: Chronic lung disease (n=4), Obstructive sleep apnoea (n=2), Type 1 Laryngeal cleft (n=2), Laryngomalacia (n=1), Other: heterogenous mixture of conditions; <sup>11</sup>These P values also show significance after applying the Bonferroni correction for multiple statistical testing.

In 21.8% of all cases with data collected, SARS-CoV-2 infection was asymptomatic. In symptomatic children, the most frequent symptom in the virtual care cohort was rhinorrhoea (117/232, 50%), and in the hospitalised cohort was cough (107/148, 72%). Fever (13/15, 87%) and cough (13/15, 87%) were the most prevalent symptoms in the small number of children requiring admission to ICU (for full description of symptom frequencies see Supplementary results).

The age-specific admission rate per 100 SARS-CoV-2 infections for medical, intensive care and PIMS-TS-specific admissions is shown in Table 1 and Figure 2. Overall, 1.38 per 100 SARS-CoV-2 infections (95% CI 1.06 to 1.46) in children aged <16 years required medical admission, and 0.09 per 100 required intensive care admission. The median length of stay for those children requiring medical admission and ward-based care was 2.0 days (IQR 1.0 to 8.0), while the median length of stay for those requiring ICU care during their admission was 7.0 days (IQR 4.0 to 11.0). The median length of stay for children admitted for social reasons was 5 days (IQR 3 to 10). There was one death associated with SARS-CoV-2 infection recorded; this child died from a confirmed invasive bacterial infection (*Streptococcus pneumoniae* meningitis). Seven children (out of 151, 4.6%) were admitted for alternative primary diagnoses and found to be incidentally SARS-CoV-2 RT-PCR positive.

**Figure 2.**
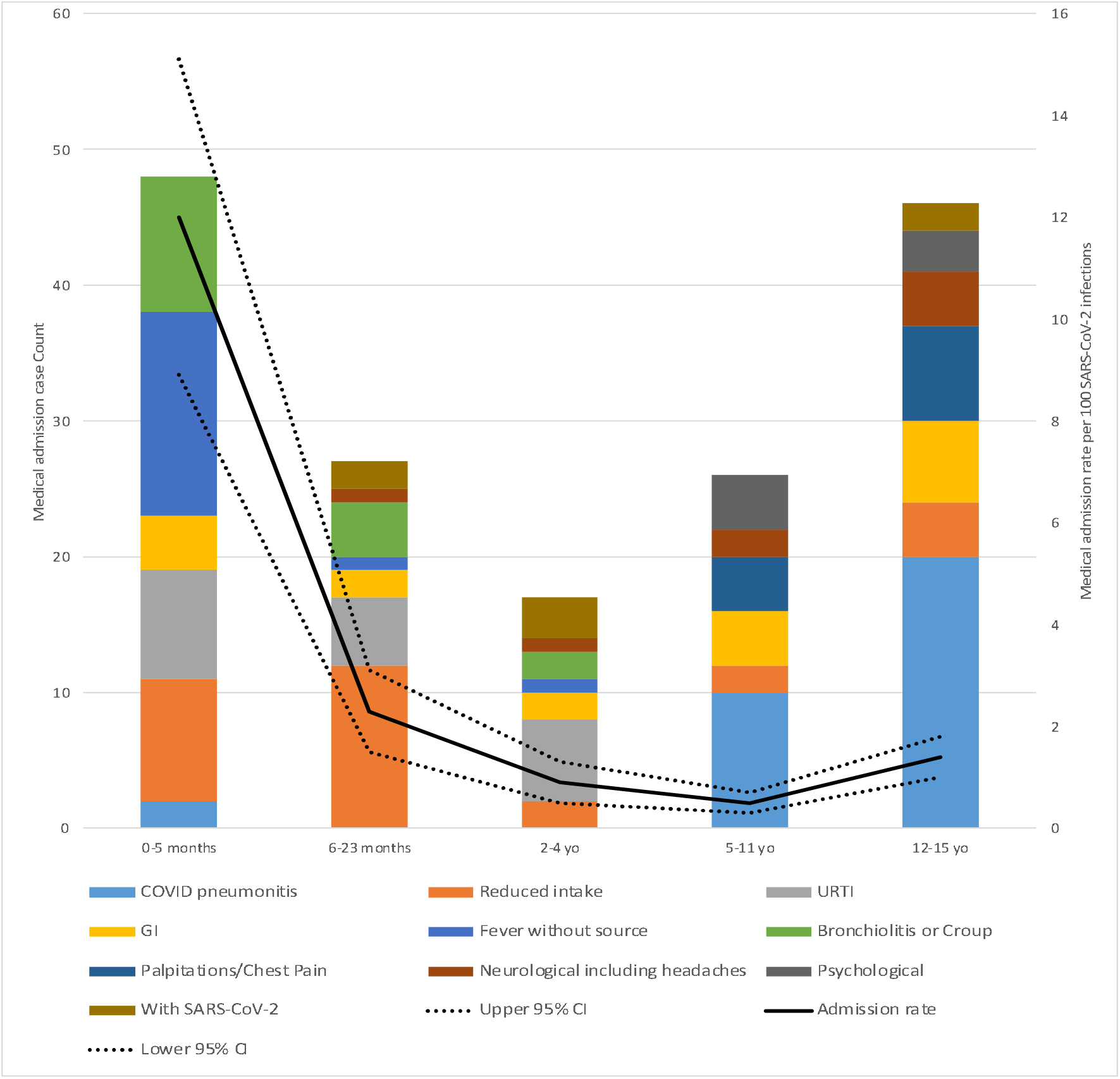
Clinician reported clinical manifestations or clinician diagnosis for age-stratified medical admissions, with age-specific admission rate per 100 SARS-CoV-2 infections. Notes: ‘COVID pneumonitis’ includes 4 cases with fever and breathlessness, but without chest X-ray performed; reduced intake includes all cases where inadequate fluid intake was the primary reason for admission; gastrointestinal (GI) included vomiting and/or diarrhoea and/or abdominal pain without a specific diagnosis; psychological included acute mental state or behavioural disturbance without an identified organic cause; palpitations/chest pain included one case of confirmed pericarditis; neurological included seizure, meningitis, encephalopathy and headaches; with SARS-CoV-2 infection included cases of pilonidal sinus, subdural empyema, trauma, toxin ingestion, bacterial lymphadenitis.

Demographics, medical co-morbidities and clinical features by severity of disease are outlined in Tables 3 and 4. The median age for those in the virtual care cohort was 7.8 years (IQR 3.8 to 11.4 years); while for medical and ICU admission was 2.0 years (IQR 0.3 to 11.2) and 12.8 years (IQR 10.4 to 15.5), respectively. Obesity (defined as a weight > 95^th^ centile) was a significant risk factor for ICU admission (57% of ICU patients vs 19% of medical admissions and 29% of outpatients, p = 0.004). A history of prematurity (defined as a gestational age at delivery of ≤36 weeks) was associated with both medical and ICU admission (14% and 15% of these admissions, respectively). Asthma and viral induced wheeze were not associated with admission, but other chronic respiratory conditions were associated with hospitalisation. In a multivariable model (Table 5), presence of a medical comorbidity was associated with an increased risk of medical admission (aOR 7.32, 95%CI 2.92 to 19.4, p<0.001). Identifying as an Aboriginal and Torres Strait Islander person was not associated with an increased risk for admission (aOR 2.25, 95%CI 0.31 to 14.6, p=0.4).

**Table 4.** Clinical features, treatment and outcomes of patients with SARS CoV-2 admitted to virtualKIDS-CORT care, medical (non-ICU) care and ICU at SCHN hospitals June - October 2021.

| Characteristic | VirtualKIDS-CORT sample, N = 344<sup>1</sup> | Medical (non-ICU) admission, N = 150<sup>1</sup> | ICU admission, N = 15<sup>1</sup> | P value<sup>2</sup> |
| --- | --- | --- | --- | --- |
| **Symptomatic** | 233 / 344 (68%) | 150 / 150 (100%) | 15 / 15 (100%) | **<0.001<sup>4</sup>** |
| Fever | 30 / 219 (14%) | 99 / 144 (69%) | 12 / 15 (80%) | **<0.001<sup>4</sup>** |
| Cough | 116 / 232 (50%) | 108 / 149 (72%) | 12 / 15 (80%) | **<0.001<sup>4</sup>** |
| Rhinorrhoea | 117 / 232 (50%) | 90 / 148 (61%) | 8 / 15 (53%) | 0.14 |
| Fatigue/malaise | 66 / 232 (28%) | 56 / 143 (39%) | 10 / 15 (67%) | **0.003<sup>4</sup>** |
| **Co-pathogen detected** | 0 / 343 (0%) | 9 / 150 (6.0%) | 2 / 15 (13%) | **<0.001<sup>4</sup>** |
| **Medication provided** |  |  |  |  |
| Antibiotics | 1 / 50 (2.0%) | 49 / 116 (42%) | 13 / 15 (87%) | **<0.001<sup>4</sup>** |
| Antivirals* | 0 / 50 (0%) | 1 / 116 (0.9%) | 8 / 15 (53%) | **<0.001<sup>4</sup>** |
| Antifungals | 0 / 50 (0%) | 1 / 115 (0.9%) | 2 / 15 (13%) | **0.019** |
| Corticosteroids | 0 / 50 (0%) | 18 / 116 (16%) | 14 / 15 (93%) | **<0.001<sup>4</sup>** |
| Systemic anticoagulation<sup>3</sup> | NA | 6 / 115 (5.2%) | 10 / 15 (67%) | **<0.001<sup>4</sup>** |
| **Respiratory support<sup>3</sup>** |  |  |  | **<0.001<sup>4</sup>** |
| None | NA | 134 / 144 (93%) | 1 / 15 (6.7%) |  |
| Low flow oxygen | NA | 7 / 144 (4.9%) | 2 / 15 (13%) |  |
| High flow oxygen | NA | 3 / 144 (2.1%) | 2 / 15 (13%) |  |
| Non-invasive ventilation | NA | 0 / 144 (0%) | 3 / 15 (20%) |  |
| Invasive ventilation | NA | 0 / 144 (0%) | 7 / 15 (47%) |  |
| **Length of stay (days)<sup>3</sup>** | NA | 2 (1, 8) | 7 (4, 11) | **0.015<sup>4</sup>** |
| **Death** | 0 / 344 (0%) | 0 / 150 (0%) | 1 / 15 (6.7%) | **0.029** |

Boldfaced P values show significance at an alpha level of 0.5; Abbreviations: VIW, Viral induced wheeze; ICU, Intensive care unit. Numerator and denominator data source is PAEDS. Denominators less than the sample size (N) are due to missing data.
\*Of antivirals administered, 6 children were prescribed remdesivir, and 3 children were prescribed acyclovir.
<sup>1</sup>n / N (%); <sup>2</sup> Fisher's exact test; Pearson's Chi-squared test; <sup>3</sup> Comparisons made between medical and ICU admissions only, not applicable treatments/variables for ambulatory care cases; <sup>4</sup>These P values also show significance after applying the Bonferroni correction for multiple statistical testing.

**Table 5.** Risk factors associated with inpatient admission (medical (non-ICU) admission and ICU admission) in univariate analysis and a multivariable model among cases of SARS CoV-2 infection.

| Predictor | Univariate analysis |  | Multivariable analysis<sup>1</sup> |  |
| --- | --- | --- | --- | --- |
|  | OR (95% CI) | P value | aOR (95% CI) | P value |
| **Age group<sup>2</sup>** |  | **<0.001** |  | **<0.001** |
| <6 months | 23.2 (11.4-50.3) |  | 71.0 (18.5-326) |  |
| 6 months to <2 years | 4.80 (2.50-9.29) |  | 10.5 (3.19-37.9) |  |
| 2 to <5 years | 1.80 (0.93-3.44) |  | 4.51 (1.28-16.8) |  |
| 5 to <12 years | 1.0 (ref) |  | 1.0 (ref) |  |
| 12 to <16 years | 3.47 (1.99-6.16) |  | 6.60 (2.14-22.2) |  |
| **Sex (M)** | 0.91 (0.63-1.33) | 0.6 | 0.71 (0.32-1.56) | 0.4 |
| **Indigenous<sup>3</sup>** | 1.45 (0.59-3.44) | 0.4 | 2.25 (0.31-14.6) | 0.4 |
| **Household contact<sup>4</sup>** | 0.86 (0.42-1.85) | 0.7 | 1.31 (0.24-6.94) | 0.8 |
| **Prematurity** | 2.45 (1.18-5.25) | 0.017 | 0.55 (0.15-2.07) | 0.4 |
| **Weight in the 95<sup>th</sup> percentile<sup>5</sup>** | 0.73 (0.47-1.14) | 0.2 | 0.76 (0.30-1.86) | 0.5 |
| **Comorbid condition** | 2.63 (1.73-4.00) | **<0.001** | 7.23 (2.92-19.4) | **<0.001** |
| Asthma/VIW<sup>6,7</sup> | 0.38 (0.18-0.78) | **0.008** | Not entered |  |
| Other respiratory disease<sup>7</sup> | 9.21 (1.61-174) | **0.009** | Not entered |  |
| Cardiac disease<sup>7</sup> | 2.19 (0.65-8.59) | 0.2 | Not entered |  |
| **Clinical features** |  |  |  |  |
| Fever | 14.6 (8.83-24.7) | **<0.001** | 21.1 (9.14-54.1) | **<0.001** |
| Cough | 2.73 (1.78-4.22) | **<0.001** | 2.89 (1.16-7.57) | **0.022** |
| Rhinorrhoea | 1.48 (0.99-2.23) | 0.056 | 0.53 (0.22-1.22) | 0.14 |
| Fatigue/malaise | 1.80 (1.18-2.77) | **0.007** | 1.40 (0.61-3.23) | 0.4 |

Boldfaced P values show significance at an alpha level of 0.5; Abbreviations: VIW, Viral induced wheeze; OR, odds ratio; aOR, adjusted odds ratio; CI, confidence interval. Numerator and denominator data source is PAEDS.
<sup>1</sup>Variables entered: age group, sex, indigenous status, household contact, prematurity, comorbid condition, fever, cough, rhinorrhoea, fatigue/malaise. Nagelkerke $R^2$ = 0.638; McFadden's pseudo $R^2$ = 0.47; Prediction percentage in null model = 67.58%, in test model = 85.11%; <sup>2</sup>Age at confirmation of positive PCR test; <sup>3</sup>Aboriginal and Torres Strait Islander child; <sup>4</sup>Defined as a confirmed case that was a household contact; <sup>5</sup>Presumed correlate of obesity obtained for Ambulatory care patients based on 28.9% population prevalence (NSW Ministry of Health, 2020); <sup>6</sup>Asthma and VIW cases combined for analysis due to low sample size of VIW (n = 4); <sup>7</sup>Excluded from multivariable analysis due to low sample size.

Children aged <2 years frequently presented with symptoms suggestive of common viral infections, with reduced oral intake, gastrointestinal symptoms and upper respiratory tract infection symptoms (URTIs) predominating, alongside ‘fever without a source’ in the cohort <6 months (Figure 2; Supplementary Table 2). Lower respiratory tract symptoms suggestive of COVID pneumonitis increased in frequency with age, with the highest prevalence in the oldest cohort (20/32, 37.5% in 12 to <16 years). Chest pain and palpitations occurred only in children aged >4 years (Figure 2).

There were 7 cases of PIMS-TS (0.04% of notified SARS-CoV-2 infections in NSW) (Table 1); equating to 1 per 2,496 infections. The median age of these children was 3.2 years (IQR 1.8; 7.5); four were female and one child had a medical comorbidity. Two children required ICU admission and there were no deaths. All cases occurred in the 3-6 weeks following confirmed or presumed (household compatible symptoms) SARS-CoV-2 infection, and cases clustered four weeks after peak infection counts.

294 children were admitted for social indications; their demographic data and reasons for admission are shown in Table 2. Over two-thirds of cases (n=202, 69%) were admitted due to COVID-19 in their parents/caregivers that resulted in a need for alternative adult supervision that could not be provided in the community. Other important reasons for admission included an inability for usual out-of-home-care services to care for vulnerable children (n=30, 10%).

## DISCUSSION

This is the first comprehensive analysis of the severity and disease spectrum of SARS-CoV-2 (Delta VOC) in non-hospitalised and hospitalised children from Australia. This study provides important estimates of hospital and ICU admission rates amongst children infected with SARS-CoV-2 and defines risk factors for hospital admission. We identified a high burden of social admissions to hospital in children with SARS-CoV-2 infection, reflecting that crude paediatric COVID-19 hospitalisation data may often overestimate the true burden of severe illness in children.

Our findings are consistent with the international evidence that a large proportion of children infected with SARS-CoV-2 are asymptomatic, and that those who do become symptomatic are likely to have a mild clinical illness, with rhinorrhoea the most common clinical feature.^6^ These trends have been consistent across the SARS-CoV-2 ancestral strain^28^ and Alpha and Delta VOCs^5,6^ from other locations; however the newly emerged Omicron variant as not yet been well characterised in children. It is likely that almost all children with COVID- 19 within our cohort were previously uninfected, given the extremely low seroprevalence of SARS-CoV-2 in Australian children at baseline; the disease phenotype presented here is therefore unlikely to have been modified by prior infection.

We did not find evidence that COVID-19 in children due to the Delta VOC is more severe than other variants, which concurs with other published studies.^5,6^ This is reassuring, and emphasises a need to carefully balance the harms of infection with those harms imposed by ongoing shielding of this age group – even when unvaccinated – that may be considered disproportionate to the disease risk itself.^29^ Why children remain less often infected and clinically affected by COVID-19 continues to a subject of intense interest in the literature and a number of theories – such as altered expression and affinity of SARS-CoV-2 binding receptors, and enhanced innate immunity within the upper airways – have been proposed.^30^

Our data shows that a history of medical comorbidities is an important risk factor for medical admission to hospital and intensive care, in keeping with previous studies.^3,5,31^ We did not identify a high risk of admission in those with a history of immunosuppression (also in keeping with previous studies);^30,32^ although an important caveat to this finding is that there were relatively few of these admissions among our cohort of >17,000 children. Due to the large number of children managed by virtual care, another limitation of our study was that we were unable to characterise all the demographic and medical co-morbid and clinical features among cases of all the children under this model of care; but selected a random sample that was likely representative of the larger cohort of children.

Our data revealed that obesity was associated with ICU admission, but this risk factor could not be evaluated in our multivariable model due to sample size. A history of asthma or viral induced wheeze was not associated with an increased risk of hospitalisation, yet other chronic respiratory conditions were - including chronic lung disease and obstructive sleep apnoea. This finding is consistent in the literature,^3,4,6^ and may be secondary to the T_H_2 predominant inflammation seen in asthma, with its associated IL-4, IL-5 and IL-13 response – the latter of which has been shown to downregulate the expression of SARS-CoV-2 host cell receptors.^33^

Our data revealed there are two dominant paediatric age cohorts at increased risk of medical admission due to SARS-CoV-2 infection. These are infants (<6 months) for whom a history of prematurity was an important risk factor for medical admission, and older children (aged 12-15 years), who constituted more than half the ICU admissions. The high admission rate in infants may represent a cautious management approach, with many infants being admitted for observation or supportive care only and a short length of stay. The typical viral syndromes prevalent in this age group are common reasons for admission to hospital with other viruses (such as. RSV, influenza or enteroviruses); medical hospitalisation in this cohort is common, and likely does not represent an unusual relative risk due to SARS-CoV-2.^34^ Future evaluation of the role of vaccination of pregnant women in conferring transplacental protection against SARS-CoV-2 may be an important consideration to reduce the risk of infection and burden of disease in this age group.^35^

By contrast, ‘adult-like’ COVID pneumonitis occurred with increasing frequency with older age, particularly in those ≥12 years. This cohort was at the greatest risk of moderate-severe disease, often requiring disease specific treatment; yet importantly, this population is now well covered by double-dose vaccination within NSW (78% as at 16^th^ December 2021),36 which may change the clinical spectrum of illness in this cohort going forward. Vaccination has now been approved for children ≥5 years in Australia, and will commence roll-out in January 2022.

A very small number of children were admitted during the post-infectious period with PIMS-TS at a similar frequency to the rate observed with the ancestral strain in Australia.^28,37^ The age distribution (median 3.2years) contrasted with overall medical admissions, but was consistent with that described elsewhere.^4^ All children with PIMS-TS had a favourable outcome in response to acute clinical treatment and the majority did not require ICU care.

Our data highlight the considerable resources needed to support children socially in the context of a pandemic. Two-thirds of the children admitted to hospital in our cohort were admitted for social rather than medical reasons, predominantly due to the hospitalisation of their own parents and carers. The ongoing traumatic effects of these episodes in children’s lives need further research. A significant number of highly vulnerable children admitted to hospital were solely admitted due to the lack of availability of their usual social services to provide care – including those in out-of-home care or supported by disability support agencies. This highlights the importance of up-scaling pandemic response plans for these services, and ensuring adequate resource allocation is provided to this important sector that cares for the most vulnerable children in society.

Our unique VirtualKIDS-CORT model of care has been resource-intensive but has likely avoided an excessive burden on emergency departments by providing parents with reassurance and telehealth support, alongside in-person, in-home clinical reviews when clinically indicated. Prior research has revealed frequent re- presentation to hospital by parents of children with SARS-CoV-2 infection who did not require admission; suggesting the ability to remotely monitor and manage children might remove this strain on health services and subvert exposure risk to other patients and healthcare workers.^8^ This model of care also eased the initial burden on primary care physicians, to support the rollout of COVID-19 vaccination programs and the management of non-COVID illnesses. However, this model is will likely prove unsustainable for the duration of the pandemic, and to minimise ongoing hospital resource reallocation, integrated primary care models are a key priority and are currently being established.

In summary, our findings reveal infection with SARS-CoV-2 in a largely unexposed population of children is predominantly an asymptomatic or mild disease, but that age (<6 months, or ≥12 years), obesity and the presence of comorbidities are risk factors for medical or ICU admission. Our data provides vital clinical information to guide further research and policy decisions, including considerations relevant to balancing the benefits and harms of population interventions to shield children – even when unvaccinated - from SARS-CoV- 2 infection.^3,4^ This research also establishes a baseline of clinical spectrum and severity data against which future waves on SARS-CoV-2 infection in Australia (including that currently evolving with the Omicron VOC) can be compared.

## Supporting information

Supplementary methods + results

## Data Availability

All data produced in the present study are available upon reasonable request to the authors

## Acknowledgements

Sydney Children’s Hospitals Network (SCHN) departments of General Medicine, Paediatric Intensive Care, Immunology, Rheumatology and Infectious Diseases and Infection Prevention and Control.

The many clinicians (nursing, medical,allied health) who contributed to the SCHN virtualKIDS COVID-19 positive outpatient response team.

NSW PAEDS surveillance nurses: Nicole Dinsmore, Nicole Kerly, Shirley Wong, Katherine Meredith and Laura Rost.

