## Supplementary methods + results for "COVID-19 in children in NSW, Australia, during the 2021 Delta outbreak: Severity and Disease spectrum"

Data sources and data use for calculation of rates.

| Data Source | Numerator | Denominator | Metric |
|---|---|---|---|
| Paediatric Active Enhanced Disease Surveillance (PAEDS) | SARS-CoV-2 associated ICU Admissions at SCHN | | COVID-19 ICU admission 'rate' |
| PAEDS | PIMS-TS Admissions at SCHN | | PIMS-TS admission 'rate'* |
| NSW Ministry of Health | | Statewide age-specific SARS-CoV-2 infection | |
| PAEDS | SARS-CoV-2 associated 'Medical' Admissions at SCHN | | COVID-19 hospital admission 'rate' |
| SCHN 'Home in the Hospital' program (Social admissions) | SARS-CoV-2 associated Social Admissions at SCHN | | COVID-19 social vulnerability 'rate' |
| SCHN VirtualKIDS-CORT | | SARS-CoV-2 associated Admissions at SCHN | |

Abbreviations: ICU=intensive care unit; NSW=New South Wales, Australia; PIMS-TS= paediatric inflammatory multi-system syndrome temporally associated with SARS-CoV-2; SCHN=Sydney Children's Hospital Network
*PIMS-TS = paediatric inflammatory multi-system syndrome temporally associated with SARS-CoV-2 infection; admission rate calculated using data cut of at 30 September, 2021 given lag time for this 'post-infectious' clinical manifestation.

**Supplementary Results:**

**Supplementary Table 1:** Case fatality and admission rates overall and by age group and severity of disease amongst cases of SARS-CoV-2 infection managed by SCHN hospitals June - October 2021.

| | **Rate per 100 infections [95%CI if applicable]** | | | | | |
|---|---|---|---|---|---|---|
| | **0-15 years** | **0-5 months** | **6-23 months** | **2-4 years** | **5-11 years** | **12-15 years** |
| **Death** (n=1) | 0.005 | - | - | - | - | 0.02 |
| **ICU Admit** (n=15) | 0.09 | 0.20 | 0.06 | - | 0.05 | 0.21 |
| **Medical Admit + ICU Admit** (n=165) | 1.38 [1.17 - 1.59] | 12.0 [8.9-15.1] | 2.3 [1.5-3.2] | 0.9 [0.5-1.3] | 0.5 [0.3-0.7] | 1.4 [1.0-1.8] |
| **PIMS-TS*** (n=7) | 0.04 | 0.20 | 0.06 | 0.09 | 0.03 | - |

*PIMS-TS=paediatric inflammatory multi-system syndrome temporally associated with SARS-CoV-2 infection; admission rate calculated using notification data cut of at 30 September, 2021 given lag time for this 'post-infectious' clinical manifestation.

**Supplementary Table 2.** Full list of clinical features, treatment and outcomes among cases of SARS-CoV-2 infection admitted to SCHN by highest acuity level of care amongst children aged 0-15 years managed through SCHN, June 1 - October 31, 2021.

| **Characteristic** | **VirtualKIDS-CORT sample, N = 344**[1] | **Medical (non-ICU) admission, N = 150**[1] | **ICU admission, N = 15**[1] | ***P* value**[2] |
|---|---|---|---|---|
| **Symptomatic** | 233 / 344 (68%) | 150 / 150 (100%) | 15 / 15 (100%) | **<0.001** |
| Fever | 30 / 219 (14%) | 99 / 144 (69%) | 12 / 15 (80%) | **<0.001** |
| Cough | 116 / 232 (50%) | 108 / 149 (72%) | 12 / 15 (80%) | **<0.001** |
| Rhinorrhoea | 117 / 232 (50%) | 90 / 148 (61%) | 8 / 15 (53%) | 0.14 |
| Ear pain | 2 / 230 (0.9%) | 1 / 131 (0.8%) | 0 / 14 (0%) | >0.9 |
| Oropharyngeal pain | 51 / 230 (22%) | 14 / 130 (11%) | 5 / 14 (36%) | **0.005** |
| Dyspnoea | 6 / 232 (2.6%) | 43 / 146 (29%) | 7 / 15 (47%) | **<0.001** |
| Wheeze | 2 / 232 (0.9%) | 6 / 145 (4.1%) | 0 / 15 (0%) | 0.10 |
| Chest pain | 1 / 230 (0.4%) | 15 / 130 (12%) | 1 / 14 (7.1%) | **<0.001** |
| Chest indraw | 0 / 232 (0%) | 6 / 146 (4.1%) | 0 / 15 (0%) | **0.005** |
| Myalgia | 31 / 230 (13%) | 11 / 128 (8.6%) | 6 / 14 (43%) | **0.004** |
| Arthralgia | 12 / 230 (5.2%) | 5 / 128 (3.9%) | 2 / 14 (14%) | 0.2 |
| Fatigue/malaise | 66 / 232 (28%) | 56 / 143 (39%) | 10 / 15 (67%) | **0.003** |
| Anosmia | 23 / 231 (10.0%) | 15 / 129 (12%) | 3 / 14 (21%) | 0.3 |
| Hypogeusia | 29 / 230 (13%) | 14 / 129 (11%) | 4 / 14 (29%) | 0.2 |
| Vomiting/nausea | 21 / 232 (9.1%) | 52 / 145 (36%) | 7 / 15 (47%) | **<0.001** |
| Headache | 70 / 230 (30%) | 32 / 129 (25%) | 8 / 14 (57%) | **0.042** |
| Diarrhoea | 23 / 232 (9.9%) | 42 / 147 (29%) | 7 / 15 (47%) | **<0.001** |
| Confusion/altered consciousness | 0 / 232 (0%) | 4 / 145 (2.8%) | 3 / 15 (20%) | **<0.001** |
| Seizure | 0 / 232 (0%) | 2 / 145 (1.4%) | 1 / 15 (6.7%) | **0.020** |
| Abdominal pain | 11 / 231 (4.8%) | 19 / 132 (14%) | 2 / 14 (14%) | **0.003** |
| Conjunctivitis | 15 / 232 (6.5%) | 11 / 145 (7.6%) | 0 / 15 (0%) | 0.7 |
| Rash | 10 / 232 (4.3%) | 6 / 145 (4.1%) | 3 / 15 (20%) | 0.055 |
| Ulcers | 1 / 232 (0.4%) | 0 / 145 (0%) | 0 / 15 (0%) | >0.9 |
| Lymphadenopathy | 0 / 232 (0%) | 3 / 145 (2.1%) | 0 / 15 (0%) | 0.11 |
| Bleeding/haemorrhage | 0 / 232 (0%) | 2 / 145 (1.4%) | 1 / 15 (6.7%) | **0.020** |
| Other[3] | 19 / 232 (8.2%) | 77 / 149 (52%) | 10 / 15 (67%) | **<0.001** |
| **Co-pathogen detected** | 0 / 343 (0%) | 9 / 150 (6.0%) | 2 / 15 (13%) | **<0.001** |
| **Medication provided** | | | | |
| Antibiotics | 1 / 50 (2.0%) | 49 / 116 (42%) | 13 / 15 (87%) | **<0.001** |
| Antivirals | 0 / 50 (0%) | 1 / 116 (0.9%) | 8 / 15 (53%) | **<0.001** |
| Antifungals | 0 / 50 (0%) | 1 / 115 (0.9%) | 2 / 15 (13%) | **0.019** |
| Corticosteroids | 0 / 50 (0%) | 18 / 116 (16%) | 14 / 15 (93%) | **<0.001** |
| Systemic anticoagulation | NA | 6 / 115 (5.2%) | 10 / 15 (67%) | **<0.001** |
| **Respiratory support** | | | | **<0.001** |
| None | NA | 134 / 144 (93%) | 1 / 15 (6.7%) | |
| Low flow oxygen | NA | 7 / 144 (4.9%) | 2 / 15 (13%) | |
| High flow oxygen | NA | 3 / 144 (2.1%) | 2 / 15 (13%) | |
| Non-invasive ventilation | NA | 0 / 144 (0%) | 3 / 15 (20%) | |
| Invasive ventilation | NA | 0 / 144 (0%) | 7 / 15 (47%) | |
| **Length of stay (days)** | NA | 2 (1, 8) | 7 (4, 11) | **0.015** |
| **Death** | 0 / 344 (0%) | 0 / 150 (0%) | 1 / 15 (6.7%) | **0.029** |

Boldfaced *P* values are significant at an alpha level of 0.05. Abbreviations: ICU, intensive care unit. Numerator and denominator data source is PAEDS. Denominators less than the sample size (N) are due to missing data.

[1]n / N (%);[2]Fisher's exact test; Pearson's Chi-squared test; [3H]heterogenous mix of symptoms.

**Supplementary Figure 1.** Proportion of children with symptoms (n ≥ 10*) among cases of acute SARS-CoV-2 infection admitted to SCHN by highest acuity level of care amongst children aged 0-15 years managed through SCHN, June 1 - October 31, 2021.

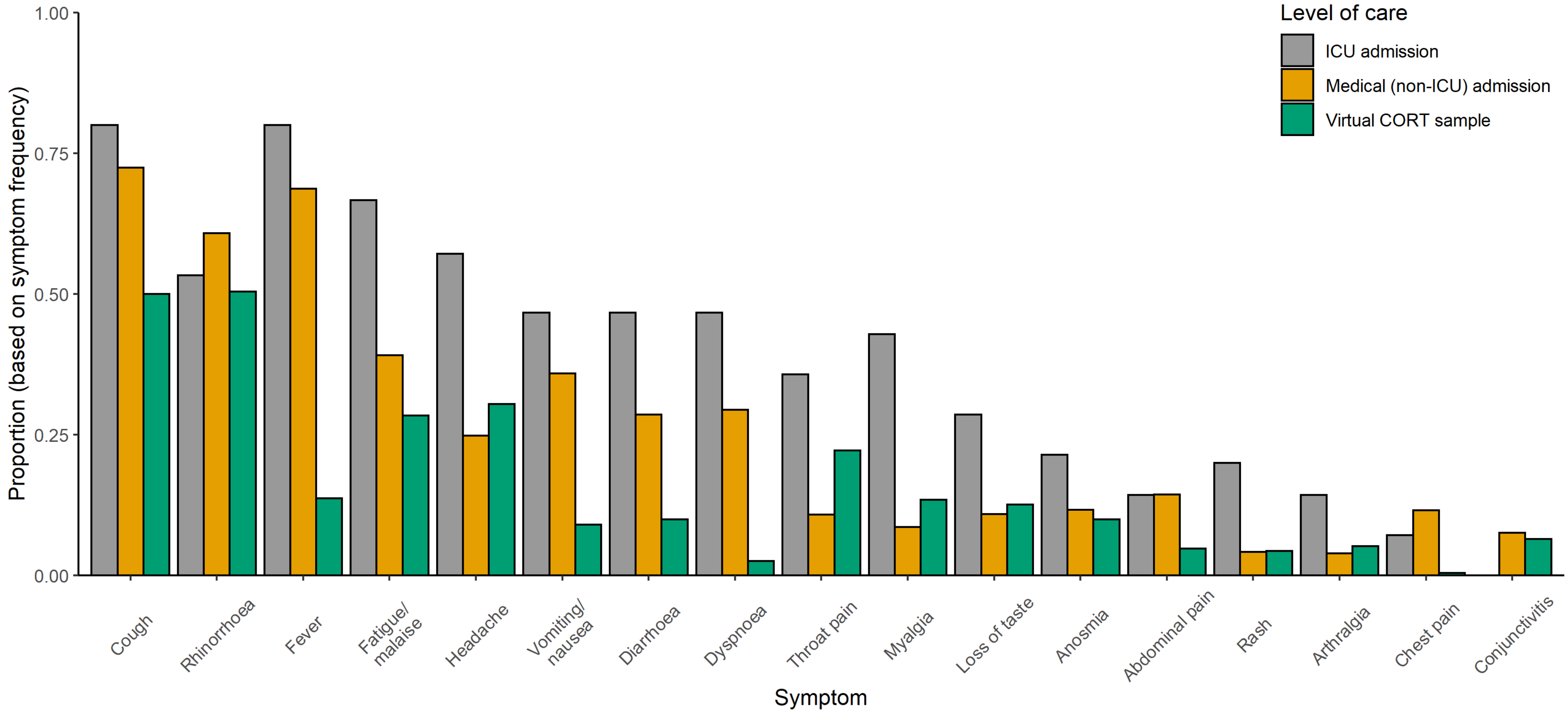


*Excluded from figure: Wheeze (n = 8), Chest indraw (n = 6), Confusion/altered consciousness (n = 7), Seizure (n = 3), Ulcer (n = 1), Lymphadenopathy (n = 3), Haemorrhage (n = 3).

**Supplementary Figure 2.** Proportion of symptoms by age group (n ≥ 10*) among cases of acute SARS-CoV-2 infection (hospitalised (non-ICU and ICU) and VirtualKIDS-CORT) managed through SCHN, June 1 - October 31, 2021.

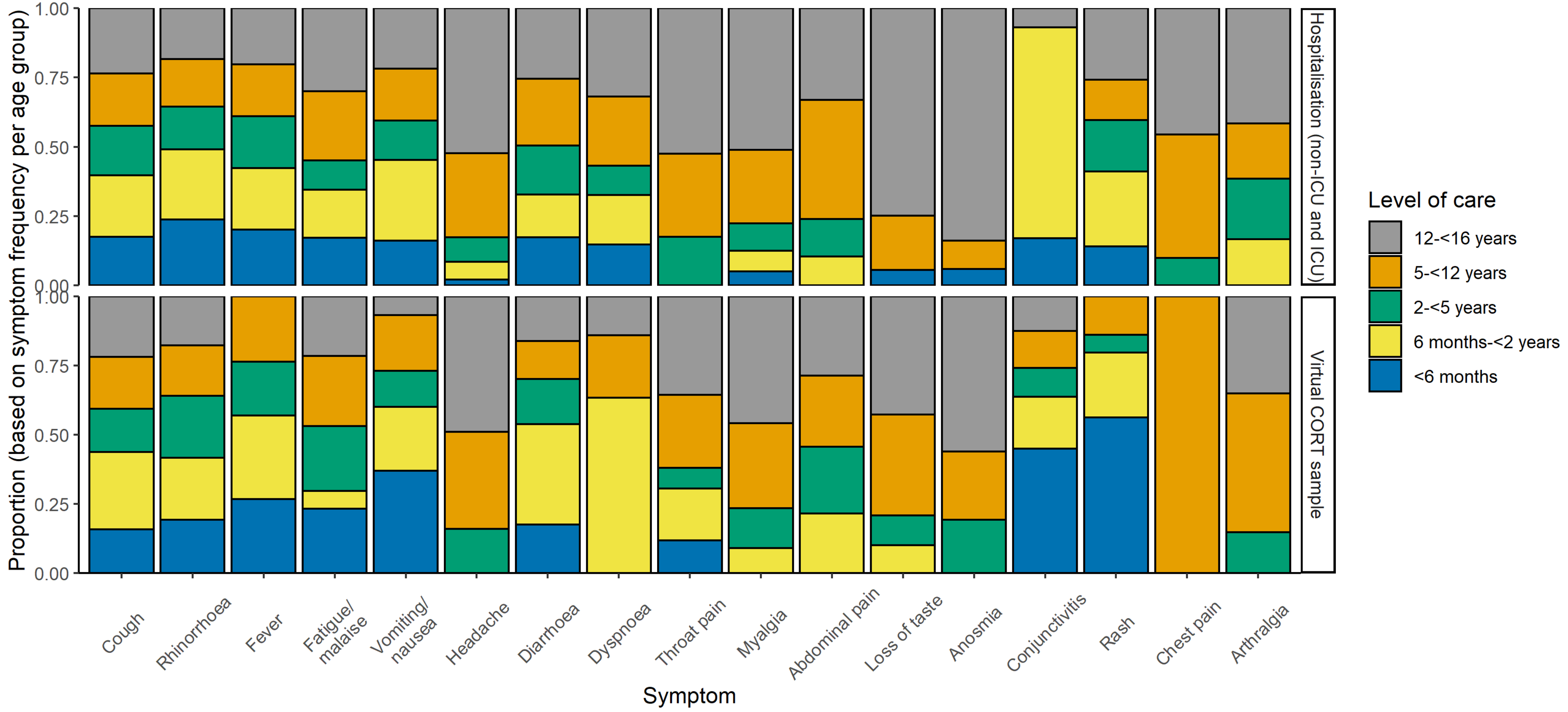


*Excluded from figure: Wheeze (n = 8), Chest indraw (n = 6), Confusion/altered consciousness (n = 7), Seizure (n = 3), Ulcer (n = 1), Lymphadenopathy (n = 3), Haemorrhage (n = 3).
